# Prioritisation of Informed Health Choices (IHC) Key Concepts to be included in lower-secondary school resources: a consensus study

**DOI:** 10.1101/2022.04.11.22273708

**Authors:** Joseph Jude Agaba, Faith Chesire, Mugisha Michael, Pamela Nandi, Jane Njue, Allen Nsangi, Venuste Nsengimana, Cyril Oyuga, Florian Rutiyomba, Daniel Semakula, Ronald Ssenyonga, Innocent Uwimana, Andrew D Oxman

## Abstract

**Background:** The Informed Health Choices Key Concepts are principles for thinking critically about healthcare claims and deciding what to do. The Key Concepts provide a framework for designing curricula, learning resources, and evaluation tools.

**Objectives:** To prioritise which of the 49 Key Concepts to include in resources for lower-secondary schools in East Africa.

**Methods:** Twelve judges used an iterative process to reach a consensus. The judges were curriculum specialists, teachers, and researchers from Kenya, Uganda, and Rwanda. After familiarising themselves with the concepts, they pilot tested draft criteria for selecting and ordering the concepts. After agreeing on the criteria, nine judges independently assessed all 49 concepts and reached an initial consensus. We sought feedback on the draft consensus from teachers and other stakeholders. After considering the feedback, nine judges independently reassessed the prioritised concepts and reached a consensus. The final set of concepts was determined after user-testing prototypes and pilot-testing the resources.

**Results:** The first panel prioritised 29 concepts. Based on feedback from teachers, students, curriculum developers, and other members of the research team, two concepts were dropped. A second panel of nine judges prioritised 17 of the 27 concepts. Due to the Covid-19 pandemic and school closures, we have only been able to develop one set of resources instead of two, as originally planned. Based on feedback on prototypes of lessons and pilot-testing a set of 10 lessons, we determined that it was possible to introduce nine concepts in 10 single-period (40 minute) lessons. We included eight of the 17 prioritised concepts and one additional concept.

**Conclusion:** Using an iterative process with explicit criteria, we prioritised nine concepts as a starting point for students to learn to think critically about healthcare claims and choices.

## Introduction

Dewey noted the importance of teaching concepts over a century ago.^1^ *“It follows that it would be impossible to overestimate the educational importance of arriving at conceptions: that is, of meanings that are* general *because applicable in a great variety of different instances in spite of their difference; that are constant, uniform, or self-identical in what they refer to, and that are standardized, known points of reference by which to get our bearings when we are plunged into the strange and unknown.”* ^2^

We have identified concepts that people need to understand and apply when deciding what to believe about health actions and what to do. “Health actions” are things that individuals or groups can do (“interventions” or “treatments”) to care for their health or the health of others. The “Informed Health Choices (IHC) Key Concepts” provide a framework for curriculum planning and designing learning resources.^3, 4^ As a first step towards developing learning resources for students in the first two years of secondary school (lower secondary school) in Kenya, Rwanda, and Uganda, we needed to decide which of the 49 IHC Key Concepts (Table 1) should be taught in this context. .*”*

**Table 1.**
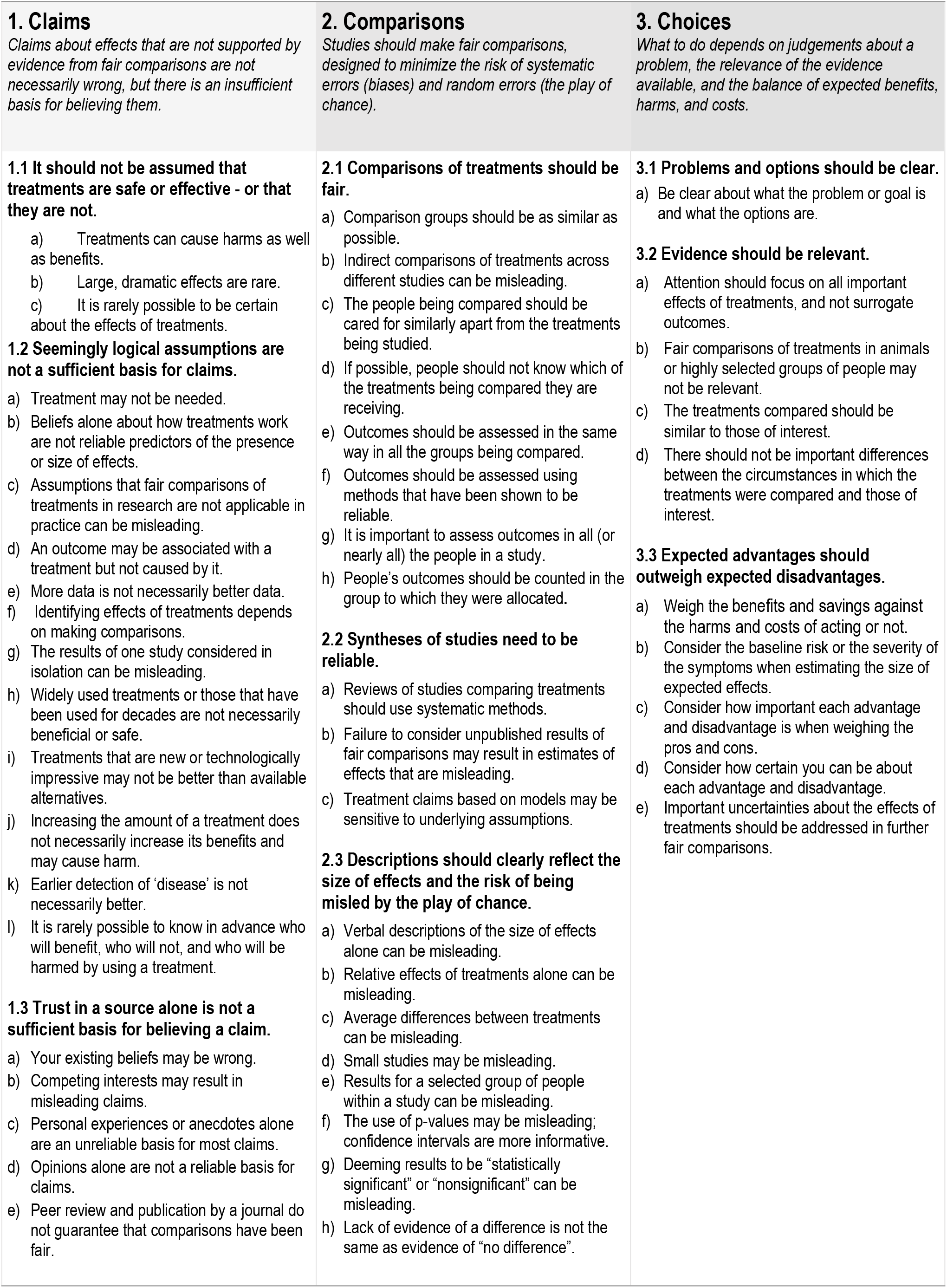
Informed Health Choices (IHC) Key Concepts

| **1. Claims**<br>*Claims about effects that are not supported by evidence from fair comparisons are not necessarily wrong, but there is an insufficient basis for believing them.* | **2. Comparisons**<br>*Studies should make fair comparisons, designed to minimize the risk of systematic errors (biases) and random errors (the play of chance).* | **3. Choices**<br>*What to do depends on judgements about a problem, the relevance of the evidence available, and the balance of expected benefits, harms, and costs.* |
| --- | --- | --- |
| **1.1 It should not be assumed that treatments are safe or effective - or that they are not.**<br>a) Treatments can cause harms as well as benefits.<br>b) Large, dramatic effects are rare.<br>c) It is rarely possible to be certain about the effects of treatments.<br>**1.2 Seemingly logical assumptions are not a sufficient basis for claims.**<br>a) Treatment may not be needed.<br>b) Beliefs alone about how treatments work are not reliable predictors of the presence or size of effects.<br>c) Assumptions that fair comparisons of treatments in research are not applicable in practice can be misleading.<br>d) An outcome may be associated with a treatment but not caused by it.<br>e) More data is not necessarily better data.<br>f) Identifying effects of treatments depends on making comparisons.<br>g) The results of one study considered in isolation can be misleading.<br>h) Widely used treatments or those that have been used for decades are not necessarily beneficial or safe.<br>i) Treatments that are new or technologically impressive may not be better than available alternatives.<br>j) Increasing the amount of a treatment does not necessarily increase its benefits and may cause harm.<br>k) Earlier detection of 'disease' is not necessarily better.<br>l) It is rarely possible to know in advance who will benefit, who will not, and who will be harmed by using a treatment.<br>**1.3 Trust in a source alone is not a sufficient basis for believing a claim.**<br>a) Your existing beliefs may be wrong.<br>b) Competing interests may result in misleading claims.<br>c) Personal experiences or anecdotes alone are an unreliable basis for most claims.<br>d) Opinions alone are not a reliable basis for claims.<br>e) Peer review and publication by a journal do not guarantee that comparisons have been fair. | **2.1 Comparisons of treatments should be fair.**<br>a) Comparison groups should be as similar as possible.<br>b) Indirect comparisons of treatments across different studies can be misleading.<br>c) The people being compared should be cared for similarly apart from the treatments being studied.<br>d) If possible, people should not know which of the treatments being compared they are receiving.<br>e) Outcomes should be assessed in the same way in all the groups being compared.<br>f) Outcomes should be assessed using methods that have been shown to be reliable.<br>g) It is important to assess outcomes in all (or nearly all) the people in a study.<br>h) People's outcomes should be counted in the group to which they were allocated.<br>**2.2 Syntheses of studies need to be reliable.**<br>a) Reviews of studies comparing treatments should use systematic methods.<br>b) Failure to consider unpublished results of fair comparisons may result in estimates of effects that are misleading.<br>c) Treatment claims based on models may be sensitive to underlying assumptions.<br>**2.3 Descriptions should clearly reflect the size of effects and the risk of being misled by the play of chance.**<br>a) Verbal descriptions of the size of effects alone can be misleading.<br>b) Relative effects of treatments alone can be misleading.<br>c) Average differences between treatments can be misleading.<br>d) Small studies may be misleading.<br>e) Results for a selected group of people within a study can be misleading.<br>f) The use of p-values may be misleading; confidence intervals are more informative.<br>g) Deeming results to be "statistically significant" or "nonsignificant" can be misleading.<br>h) Lack of evidence of a difference is not the same as evidence of "no difference". | **3.1 Problems and options should be clear.**<br>a) Be clear about what the problem or goal is and what the options are.<br>**3.2 Evidence should be relevant.**<br>a) Attention should focus on all important effects of treatments, and not surrogate outcomes.<br>b) Fair comparisons of treatments in animals or highly selected groups of people may not be relevant.<br>c) The treatments compared should be similar to those of interest.<br>d) There should not be important differences between the circumstances in which the treatments were compared and those of interest.<br>**3.3 Expected advantages should outweigh expected disadvantages.**<br>a) Weigh the benefits and savings against the harms and costs of acting or not.<br>b) Consider the baseline risk or the severity of the symptoms when estimating the size of expected effects.<br>c) Consider how important each advantage and disadvantage is when weighing the pros and cons.<br>d) Consider how certain you can be about each advantage and disadvantage.<br>e) Important uncertainties about the effects of treatments should be addressed in further fair comparisons. |

Teachers may be overwhelmed by the amount of content they are expected to cover, especially when standards are viewed as discrete and disconnected. Marzano and Kendall reviewed 160 national and state-level documents listing standards in various subject areas in the USA and synthesized the material to avoid duplication. They identified 255 content standards and 3968 discrete benchmarks that delineate what students should know and be able to do.^5^ They estimated that if teachers devoted 30 minutes of instructional time to teach each benchmark, they would need an additional 15,465 hours (nine school years).

This is consistent with findings of a process evaluation we conducted to explore barriers to scaling up use of the IHC primary school intervention in Uganda.^6^ The intervention consisted of providing the IHC primary school resources, as well as teacher training workshops. It was shown to have a large effect on primary school children’s ability to think critically about health claims,^7^ which was sustained after one year.^8^ Teachers who used the primary school intervention in the trial said: the IHC Key Concepts were important; they were motivated to teach the concepts; and the children were enthusiastic about the lessons. The main barrier we identified to scaling up use of the intervention was the need to incorporate the lessons in the national curriculum. The IHC lessons were viewed as an addition to what was already a packed primary school curriculum.

It is essential to prioritise what to include in school curricula. Wiggins and McTighe argue that prioritising should focus on “big ideas” and “core tasks”; “A big idea is a concept, theme, or issue that gives meaning and connection to discrete facts and skills,” while a core task is “the most important performance demands in any field”.^9^ Priorities should be established by building upon the big ideas and by focusing schoolwork around core tasks or “transfer tasks” derived from authentic challenges. In the same vein, Bruner writes: *“For any subject taught in* […] *school, we might ask* [is it] *worth an adult’s knowing, and whether having known it as a child makes a person a better adult. A negative or ambiguous answer means the material is cluttering up the curriculum.”*^10^

Bruner’s idea of a “spiral curriculum” is based on recurring, deepening inquiries into big ideas and important tasks, helping students learn in a way that is developmentally sensible and effective; *“The basic ideas at the heart of all science and mathematics and the basic themes that give form to life and literature are as simple as they are powerful. To be in command of these basic ideas and use them effectively requires a continual deepening of one’s understanding of them that comes from learning to use them in progressively more complex forms*The basic principle underlying Wiggins’ and McTighe’s approach to curriculum design - “backward design” - is to begin with the desired, final outcomes and to focus on the learner’s needs. Rather than building a curriculum around the logic of the content, it should be designed around the needs of learners trying to understand the big ideas and to perform the core tasks.

This paper describes a process in line with the thinking of Wiggins and McTighe and of Bruner, in which we prioritised and ordered IHC Key Concepts to be included in lower-secondary school resources for Kenya, Rwanda, and Uganda. Our intention was to develop a spiral curriculum for a series of resources. However, due to the Covid-19 pandemic, school closures, and project delays, we only developed one set of resources (for a single school term) and prioritised IHC Key Concepts for inclusion in that set of resources.

### Objective

To prioritise and order IHC Key Concepts to be included in the IHC lower-secondary school resources.

## Methods

We used an iterative, structured consensus process built on Wiggins’ and McTighe’s “backward design” approach,^9^ the Nominal Group Technique consensus process,^11^ and Feinstein’s criteria for sensibility.^12^ The process included the following steps:

1. Selecting and training the judges
2. Deciding on criteria and response options for judgements
3. Prioritising and ordering concepts
4. Reaching initial consensus
5. Collecting feedback
6. Prioritising and ordering concepts and reaching final consensus

Ten judges initially prioritised the IHC Key Concepts to be included in the IHC secondary school resources (Table 2). Three were curriculum specialists or teachers, one was a health promotion officer, and the other six were health researchers who were members of the project team and were familiar with the IHC Key Concepts.

**Table 2.** Judges for the initial prioritisation*

| Country | Organisation | Background |
| --- | --- | --- |
| Kenya | Huma Girls Secondary School | Teacher |
| Kenya | Kenya Institute of Curriculum Development | Curriculum specialist |
| Kenya | Kisumu County | Health promotion officer |
| Kenya | Tropical Institute of Community Health and Development | Researcher, health policy and public health |
| Rwanda | University of Rwanda, College of Medicine and Health Sciences | Researcher, public health |
| Rwanda | University of Rwanda, College of Medicine and Health Sciences | Researcher, public health |
| Uganda | Makerere University, College of Health Sciences | Physician, researcher |
| Uganda | Makerere University, College of Health Sciences | Researcher |
| Uganda | Makerere University, College of Health Sciences | Researcher, Clinical epidemiology and biostatistics |

\* These nine judges completed an independent assessment of the Key Concepts using the criteria in Table 3. A curriculum specialist from the Ugandan National Curriculum Development Centre participated in discussions but did not complete an independent assessment of the Key Concepts.

At the first online meeting of the panel in November 2019, ADO presented an overview of the research project and introduced the IHC Key Concepts, the protocol for prioritising the concepts, and plans for pilot testing proposed criteria for prioritising the concepts.^13^ Three criteria were used in the pilot to make two judgements about each of nine Key Concepts:

### Criteria

- Are the learners likely to be able to understand and use the concept?
- How important is the concept to the learners?
- Are there sufficient time and resources to help them learn the concept?

### Judgements

- Should the concept be included?
- When should it be included?

Following the pilot, we agreed on six revised criteria (Table 3). These included the importance of each concept for the “central ideas” and the “core tasks” (Table 4). Nine of the judges then independently assessed each of the 49 Key Concepts using those criteria. ADO summarised their assessments and facilitated a discussion that resulted in an initial prioritisation of the Key Concepts. During the discussion, people from each end of the range of judgements for each concept were invited to provide the reasons for their judgements, before others were invited to comment. The outcome of the meeting was summarised, fed back to the judges, and agreed.

**Table 3.**
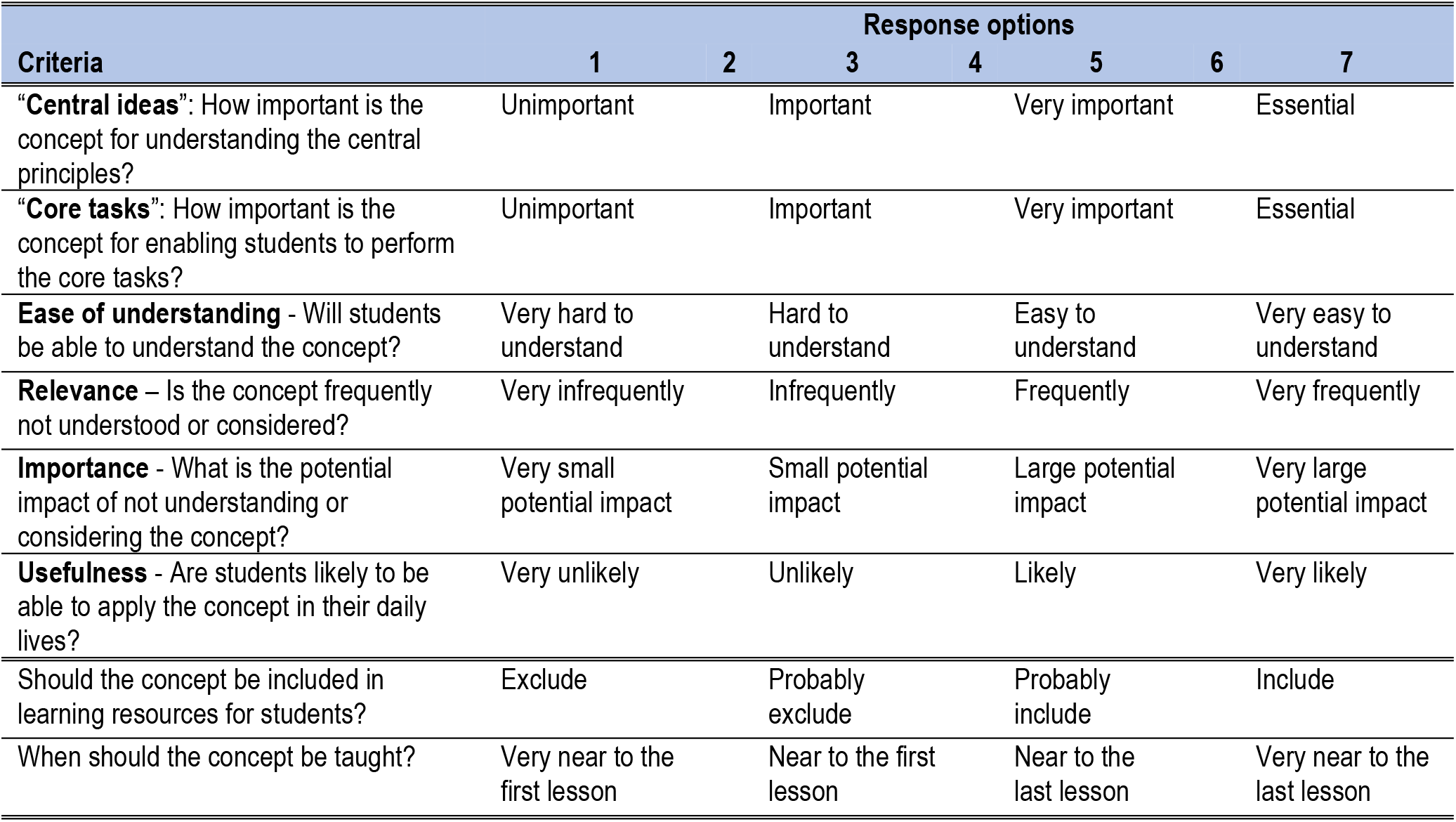
Criteria and response options for judgements

| Criteria | Response options |  |  |  |  |  |  |
| --- | --- | --- | --- | --- | --- | --- | --- |
|  | 1 | 2 | 3 | 4 | 5 | 6 | 7 |
| “**Central ideas**”: How important is the concept for understanding the central principles? | Unimportant |  | Important |  | Very important |  | Essential |
| “**Core tasks**”: How important is the concept for enabling students to perform the core tasks? | Unimportant |  | Important |  | Very important |  | Essential |
| **Ease of understanding** - Will students be able to understand the concept? | Very hard to understand |  | Hard to understand |  | Easy to understand |  | Very easy to understand |
| **Relevance** – Is the concept frequently not understood or considered? | Very infrequently |  | Infrequently |  | Frequently |  | Very frequently |
| **Importance** - What is the potential impact of not understanding or considering the concept? | Very small potential impact |  | Small potential impact |  | Large potential impact |  | Very large potential impact |
| **Usefulness** - Are students likely to be able to apply the concept in their daily lives? | Very unlikely |  | Unlikely |  | Likely |  | Very likely |
| Should the concept be included in learning resources for students? | Exclude |  | Probably exclude |  | Probably include |  | Include |
| When should the concept be taught? | Very near to the first lesson |  | Near to the first lesson |  | Near to the last lesson |  | Very near to the last lesson |

**Table 4.** Central ideas and core tasks

| “Central ideas” and “core abilities” |
| --- |
| **Central ideas** are central principles* underlying what students should learn.<br>**Core abilities** are essential actions that students should be able to perform. |
| **Central idea 1:** Claims about effects should be supported by evidence from fair comparisons.<br>*NB: Other claims are not necessarily wrong, but there is an insufficient basis for believing them.*<br>• **Core ability 1:** Recognise when a claim has an untrustworthy basis<br>*I.e. Recognise claims about effects; that claims should be questioned; when a claim has an untrustworthy basis; and when to think more carefully about claims* |
| **Central idea 2:** Evidence of effects should come from fair comparisons, designed to minimize the risk of systematic errors (biases) and random errors (the play of chance).<br>• **Core ability 2:** When there is evidence used to support a claim about effects, recognise whether it is trustworthy or untrustworthy<br>*I.e. Whether comparisons are fair or unfair, whether summaries of comparisons (reviews) are reliable or unreliable, and when presentations of effects are helpful or misleading* |
| **Central idea 3:** Good choices depend on using the best available information.<br>*I.e. Information regarding: the problem and options; the relevance of the best available evidence on the effects of those options; and the balance of expected benefits, harm, and costs*<br>• **Core ability 3:** Make informed choices to achieve health outcomes and other goals<br>*I.e. Clarifying and understanding the problem and options when making decisions about what to do; judging the relevance of evidence used to inform those decisions; weighing the advantages and disadvantages of the options, taking into account how important the benefits and harms are, the costs, and the certainty of the evidence; and communicating with others about the advantages and disadvantages of the options* |

\* Fundamental truths or propositions that serve as the foundation

We collected informal feedback on the initial prioritisation from teacher and student networks and advisory groups in each country, from our international advisory group, and from other members of our research team.^14^ An introductory meeting was held with a second panel of judges (Table 5). The second panel assessed the Key Concepts from the first prioritisation. ADO summarised their assessments and facilitated a discussion that resulted in a final set of prioritised Key Concepts. The final set of Key Concepts that are included in the IHC secondary school resources was decided by the research team following user feedback on prototypes of the resources.

**Table 5.** Judges for the final prioritisation*

| Country | Organisation | Background |
| --- | --- | --- |
| Kenya † | Huma Girls Secondary School | Teacher |
| Kenya † | Kenya Institute of Curriculum Development | Curriculum specialist |
| Kenya † | Tropical Institute of Community Health and Development | Researcher, health policy and public health |
| Rwanda | Rwanda Education Board | Curriculum specialist |
| Rwanda | University of Rwanda, College of Education | Researcher, education |
| Rwanda † | University of Rwanda, College of Medicine and Health Sciences | Researcher, public health |
| Uganda † | Makerere University, College of Health Sciences | Physician, researcher |
| Uganda † | Makerere University, College of Health Sciences | Researcher |
| Uganda † | Makerere University, College of Health Sciences | Researcher, Clinical epidemiology and biostatistics |

\* These nine judges completed an independent assessment of the Key Concepts using the criteria in Table 3. A second curriculum specialist from the Kenya Institute of Curriculum Development, a second curriculum specialist from the Rwanda Education Board, and a curriculum specialist from the Ugandan National Curriculum Development Centre participated in discussions but did not complete an independent assessment of the Key Concepts.
† Also participated in the initial consensus and completed an independent assessment of all 49 Key Concepts.

In addition to prioritising the concepts, the panels also made judgements about when each included concept should be taught (Table 3).

## Results

In the initial prioritisation, the average score for including each of the 49 concepts ranged from seven (for “Treatments can cause harms as well as benefits,” “Your existing beliefs may be wrong,” and “Personal experiences or anecdotes alone are an unreliable basis for most claims.”) to 1.2 (for “Failure to consider unpublished results of fair comparisons may result in estimates of effects that are misleading” and “Treatment claims based on models may be sensitive to underlying assumptions.”) (Supporting information 1). Seventeen concepts had an average score greater than six. Nine concepts had an average score between five and six, eight had an average score between four and five, and 15 had an average score less than four. After discussing the scores, the panel agreed on prioritising 29 of the 49 concepts (Table 6).

**Table 6.** Prioritised IHC concepts

| IHC Key Concepts | Consensus |  |  |  |
| --- | --- | --- | --- | --- |
|  | 1 | 2a | 2b | Final |
| **Claims** |  |  |  |  |
| **It should not be assumed that treatments are safe or effective - or that they are not.** |  |  |  |  |
| 1. Treatments can cause harms as well as benefits. | ✓ | ✓ | ✓ | ✓ |
| 2. Large, dramatic effects are rare. | ✓ | ✓ | ✓ | ✓ |
| 3. It is rarely possible to be certain about the effects of treatments. | ✓ | ✓ |  |  |
| **Seemingly logical assumptions are not a sufficient basis for claims.** |  |  |  |  |
| 4. Treatment may not be needed. | ✓ | ✓ |  |  |
| 5. Beliefs alone about how treatments work are not reliable predictors of the presence or size of effects. | ✓ | ✓ | ✓ |  |
| 6. Assumptions that fair comparisons of treatments in research are not applicable in practice can be misleading. |  |  |  |  |
| 7. An outcome may be associated with a treatment but not caused by it. | ✓ | ✓ | ✓ |  |
| 8. More data is not necessarily better data. |  |  |  |  |
| 9. Identifying effects of treatments depends on making comparisons. | ✓ | ✓ | ✓ | ✓ |
| 10. The results of one study considered in isolation can be misleading. | ✓ | ✓ |  |  |
| 11. Widely used treatments or those that have been used for decades are not necessarily beneficial or safe. | ✓ | ✓ | ✓ | ✓ |
| 12. Treatments that are new or technologically impressive may not be better than available alternatives. | ✓ | ✓ | ✓ | ✓ |
| 13. Increasing the amount of a treatment does not necessarily increase its benefits and may cause harm. | ✓ | ✓ | ✓ |  |
| 14. Earlier detection of 'disease' is not necessarily better. |  |  |  |  |
| 15. It is rarely possible to know in advance who will benefit, who will not, and who will be harmed by using a treatment. |  |  |  |  |
| **Trust in a source alone is not a sufficient basis for believing a claim.** |  |  |  |  |
| 16. Your existing beliefs may be wrong. | ✓ | ✓ | ✓ |  |
| 17. Competing interests may result in misleading claims. | ✓ | ✓ | ✓ |  |
| 18. Personal experiences or anecdotes alone are an unreliable basis for most claims. | ✓ | ✓ | ✓ | ✓ |
| 19. Opinions alone are not a reliable basis for claims. | ✓ | ✓ | ✓ |  |
| 20. Peer review and publication by a journal do not guarantee that comparisons have been fair. |  |  |  |  |
| **Comparison** |  |  |  |  |
| **Comparisons of treatments should be fair.** |  |  |  |  |
| 21. Comparison groups should be as similar as possible. | ✓ | ✓ | ✓ | ✓ |
| 22. Indirect comparisons of treatments across different studies can be misleading. |  |  |  |  |
| 23. The people being compared should be cared for similarly apart from the treatments being studied. | ✓ | ✓ | ✓ |  |
| 24. If possible, people should not know which of the treatments being compared they are receiving. | ✓ |  |  |  |
| 25. Outcomes should be assessed in the same way in all the groups being compared. | ✓ | ✓ | ✓ |  |
| 26. Outcomes should be assessed using methods that have been shown to be reliable. |  |  |  |  |
| 27. It is important to assess outcomes in all (or nearly all) the people in a study. |  |  |  |  |
| 28. People's outcomes should be counted in the group to which they were allocated. |  |  |  |  |
| **Syntheses of studies need to be reliable.** |  |  |  |  |
| 29. Reviews of studies comparing treatments should use systematic methods. |  |  |  |  |
| 30. Failure to consider unpublished results of fair comparisons may result in estimates of effects that are misleading. |  |  |  |  |
| 31. Treatment claims based on models may be sensitive to underlying assumptions. |  |  |  |  |
| **Descriptions should clearly reflect the size of effects and the risk of being misled by the play of chance.** |  |  |  |  |
|  | 1 | 2a | 2b | Final |
| 32. Verbal descriptions of the size of effects alone can be misleading. | ✓ | ✓ |  |  |
| 33. Relative effects of treatments alone can be misleading. | ✓ | ✓ |  |  |
| 34. Average differences between treatments can be misleading. | ✓ |  |  |  |
| 35. Small studies may be misleading. | ✓ | ✓ |  | ✓ |
| 36. Results for a selected group of people within a study can be misleading. |  |  |  |  |
| 37. The use of p-values may be misleading; confidence intervals are more informative. |  |  |  |  |
| 38. Deeming results to be “statistically significant” or “nonsignificant” can be misleading. |  |  |  |  |
| 39. Lack of evidence of a difference is not the same as evidence of “no difference”. |  |  |  |  |
| **Choices** |  |  |  |  |
| **Problems and options should be clear.** |  |  |  |  |
| 40. Be clear about what the problem or goal is and what the options are. | ✓ | ✓ | ✓ |  |
| **Evidence should be relevant.** |  |  |  |  |
| 41. Attention should focus on all important effects of treatments, and not surrogate outcomes. |  |  |  |  |
| 42. Fair comparisons of treatments in animals or highly selected groups of people may not be relevant. | ✓ | ✓ |  |  |
| 43. The treatments compared should be similar to those of interest. |  |  |  |  |
| 44. There should not be important differences between the circumstances in which the treatments were compared and those of interest. |  |  |  |  |
| **Expected advantages should outweigh expected disadvantages.** |  |  |  |  |
| 45. Weigh the benefits and savings against the harms and costs of acting or not. | ✓ | ✓ | ✓ | ✓ |
| 46. Consider the baseline risk or the severity of the symptoms when estimating the size of expected effects. | ✓ | ✓ |  |  |
| 47. Consider how important each advantage and disadvantage is when weighing the pros and cons. | ✓ | ✓ |  |  |
| 48. Consider how certain you can be about each advantage and disadvantage. | ✓ | ✓ |  |  |
| 49. Important uncertainties about the effects of treatments should be addressed in further fair comparisons. |  |  |  |  |
| **Number of concepts** | 29 | 27 | 17 | 9 |

\* ✓ = Included
1 = First consensus
2a = Concepts assessed by the second consensus panel. Two concepts prioritised by the first panel “If possible, people should not know which of the treatments being compared they are receiving.” and “Average differences between treatments can be misleading.” were not considered after feedback from teachers, students, curriculum developers, and other members of the research team.
2b = Concepts prioritised by the second consensus panel
Final = Prioritised concepts after collecting feedback on prototypes of the learning resources and agreed on by the second consensus panel. One concept that was not initially prioritised by the second consensus panel (“Small studies may be misleading.”) was included as one of the nine IHC Key Concepts included in the secondary school resources.

Two of the 29 concepts were not included in the feedback from teachers, students, curriculum developers, and other members of the research team.: “If possible, people should not know which of the treatments being compared they are receiving” and “Average differences between treatments can be misleading.” (Table 6).

The average score for the 27 concepts ranged from seven (for “Treatments can cause harms as well as benefits.”) to 4.3 (for “The results of one study considered in isolation can be misleading.”) (Supporting information 2). After discussing the scores, the panel agreed on prioritising 17 of the Key Concepts.

Our original plan was to develop two sets of learning resources to be used during two school terms. However, due to the Covid-19 pandemic, school closures, and project delays, it was only possible to produce one set of resources for a single school term. After collecting feedback on prototypes of the learning resources, the second consensus panel agreed on the nine concepts included in the IHC secondary school resources (Table 6).

One concept that was not one of the 17 prioritised concepts, was included as one of the nine concepts: “Small studies may be misleading.” That concept had the same average score (5.4) as the two included concepts with the lowest score. Only one other concept was included from the second group of concepts (about comparisons): “Comparison groups should be as similar as possible.” It was decided that it was important to include the additional concept and consideration of the risk of being misled by the play of chance, together with consideration of the risk of bias.

### Ordering of the concepts

Based on the average score for the nine judges in the second panel, the prioritised concepts were ordered as shown in Table 7. However, based on feedback on early prototypes of the lessons and pilot testing a complete version of the lessons, the order in which the concepts are introduced was modified. The final order in which the concepts were introduced in the 10 lessons is shown in the last column in Table 7, and an overview of the 10 lessons is shown in Table 8. Because students and teachers, as well as others, frequently understand “treatment” narrowly to only include medical care given to a patient, we have used “health action” in the secondary school resources.

**Table 7.** Ordering of the concepts

| Concept | Average judgement | Lesson |
| --- | --- | --- |
| 1. Treatments can cause harms as well as benefits. | Very near to the first lesson | 1,2 |
| 2. Widely used treatments or those that have been used for decades are not necessarily beneficial or safe. | Very near to the first lesson | 3 |
| 3. Treatments that are new or technologically impressive may not be better than available alternatives. | Near to the first lesson | 3 |
| 4. Personal experiences or anecdotes alone are an unreliable basis for most claims. | Near to the first lesson | 3 |
| 5. Beliefs alone about how treatments work are not reliable predictors of the presence or size of effects. | Near to the first lesson |  |
| 6. Increasing the amount of a treatment does not necessarily increase its benefits and may cause harm. | Near to the first lesson |  |
| 7. Your existing beliefs may be wrong. | Near to the first lesson |  |
| 8. Opinions alone are not a reliable basis for claims. | Near to the first lesson |  |
| 9. Competing interests may result in misleading claims. | Near to the last lesson |  |
| 10. An outcome may be associated with a treatment but not caused by it. | Near to the last lesson |  |
| 11. Comparison groups should be as similar as possible. | Near to the last lesson | 6 |
| 12. Outcomes should be assessed in the same way in all the groups being compared. | Near to the last lesson |  |
| 13. Be clear about what the problem or goal is and what the options are. | Near to the last lesson |  |
| 14. Identifying effects of treatments depends on making comparisons. | Very near to the last lesson | 4 |
| 15. Large, dramatic effects are rare. | Very near to the last lesson | 3 |
| 16. The people being compared should be cared for similarly apart from the treatments being studied. | Very near to the last lesson |  |
| 17. Weigh the benefits and savings against the harms and costs of acting or not. | Very near to the last lesson | 8,9 |
| 18. Small studies may be misleading. | Very near the last lesson | 7 |

**Table 8.** Overview of the lessons

| Lesson | Learning goals and concepts |
| --- | --- |
| **Part 1. Claims about effects that are not supported by reliable comparisons are not necessarily wrong, but there is a weak basis for believing them.**<br>*The first six concepts can help people to recognise when a claim about a health action has a weak basis. By “claim”, we mean something that someone says as if it is true, but it may be false. “Claim” has more than one meaning. In these resources, it means a statement of a belief or opinion about something. The focus is on claims about the effects of doing something; specifically, claims about the effects of health actions.* |  |
| **1. Health actions** | By the end of this lesson, students should be able to identify health actions, and explain why it is important to think critically about health actions. |
|  | **Concept**<br>➤ *Health actions can have helpful effects, but they can also have harmful effects and be expensive.* |
| **2. Health claims** | By the end of this lesson, students should be able to identify claims about the effects of health actions and their three main parts, and explain why it is important to think critically about such claims. |
|  | **Concept**<br>➤ *Health actions can have helpful effects, but they can also have harmful effects and be expensive. People often exaggerate the benefits of treatments and ignore or downplay potential harms. However, few effective treatments are 100% safe.* |
| **3. Unreliable claims** | By the end of this lesson, students should be able to identify claims about the effects of health actions that are only based on personal experiences, how commonly-used or for how long something has been used, or how new or expensive something is, and explain why most such claims are unreliable. |
|  | **Concepts**<br>➤ *Most health actions do not have obvious effects that occur shortly after the action and where everyone or nearly everyone experiences the same effect.*<br>➤ *Usually, personal experience (something that happened to someone after taking a health action) is a weak basis for claims about the effects of health actions. The problem is that we cannot know what would have happened without the health action.*<br>➤ *Health actions that have not been evaluated in a reliable comparison but are commonly used or have been used for a long time are often assumed to work. However, they might not work and might be harmful or wasteful.*<br>➤ *Similarly, health actions that have not been evaluated in a reliable comparison but are new, expensive, or technologically impressive are often assumed to work. However, they also might not work and might be harmful or wasteful.* |
| **4. Reliable claims** | By the end of this lesson, students should be able to explain why knowledge about the effects of health actions depends on comparisons, and explain why we need researchers to make the comparisons. |
|  | **Concept**<br>➤ *Knowledge about the effects of health actions depends on comparisons. Unless a health action is compared to something else, it is not possible to know what would happen without the health action. Reliable comparisons require special knowledge and skills that health researchers have, as well as a lot of resources.* |
| **5. Using what we learned (1)** | By the end of this lesson, students should be able to remember what they learned in Lessons 1 to 4, apply what they have learned in their daily lives, and recognise limits to what they have learned. |
| **Part 2. Comparisons between health actions should be reliable.**<br>*These resources focus on two concepts that can help people to recognise reliable comparisons between health actions and to avoid being misled by unreliable comparisons between health actions.* |  |
| **6. Randomly-created groups** | By the end of this lesson, students should be able to explain why groups of people in a comparison should be similar at the start, and why researchers should randomly create groups, when possible, to create similar groups. |
|  | **Concept**<br>► In a comparison between health actions, important differences between comparison groups might result in the effects appearing either larger or smaller than they actually are. Randomly creating groups makes sure groups of people are as similar as possible at the start of a comparison and avoids unknown differences. For this reason, the most reliable evidence comes from comparisons of randomly-created groups (randomised trials). |
| **7. Large-enough groups** | By the end of this lesson, students should be able to explain what it means for comparisons between health actions to be large enough. |
|  | **Concept**<br>► If a comparison between health actions is too small, we cannot be sure that the results reflect a true difference (or lack of difference) between the effects of the different health actions. The results could just be by chance. |
| **Part 3. Making a smart choice about a problem depends on making judgements about evidence from reliable comparisons between health actions.**<br>*These resources focus on one concept that can help people make smart choices.* |  |
| **8. Personal choices** | By the end of this lesson, students should be able to identify some advantages and disadvantages of health actions, for individuals, and explain the importance of weighing the advantages of a health action against the disadvantages. |
|  | **Concept**<br>► People making a personal choice about whether to take a health action should consider the potential benefits and potential harms, costs, other advantages and disadvantages, and other alternatives. |
| **9. Community choices** | By the end of this lesson, students should be able to identify advantages and disadvantages of health actions for communities, and explain the importance of weighing the advantages of a community health action against the disadvantages. |
|  | **Concept**<br>► People making a community choice about whether to take a health action should consider the potential benefits and potential harms, costs, other advantages and disadvantages, and other alternatives. They should also consider who will benefit, who will be harmed, who will save resources, who will pay, and whether it is fair. |
| **10. Using what we learned (2)** | By the end of this lesson, students should be able to remember what they learned in Lessons 1 to 9, apply what they have learned in their daily lives, and recognise limits to what they have learned. |

## Discussion

Using an iterative process that engaged curriculum planners and teachers as well as researchers, we prioritised 17 key concepts to teach lower secondary school students in Kenya, Rwanda, and Uganda. The prioritised and ordered Key Concepts were the starting point for the IHC secondary school resources that we developed for lower-secondary schools in East Africa. In parallel, we conducted a context analysis in each country. The context analyses explored where teaching the Key Concepts best fits in the curriculum. They also explored conditions for introducing the learning resources into schools, such as the availability of time, the availability and use of digital learning resources, who decides what learning resources are used and how, and what influences their decisions.

Based on feedback on prototypes of the lessons and due to the Covid-19 pandemic and school closures, we prioritised eight of the concepts and one concept not prioritised by the second consensus panel. There were several reasons for this. First, because of limitations to our funding and the pandemic, we decided only to develop a single set of lessons that could be taught in a single school term, instead of two sets of learning resources. Based on feedback from curriculum specialists and teachers in the three countries, we determined that it would be difficult to find time in the curriculum for more than 10 single period (40-minute) lessons. Moreover, based on feedback on prototypes of the lessons, we determined that it was necessary to introduce some basic terminology in the first two lessons, such as health action, effect, and claim. In addition, the last lesson was used as a review of the first nine lessons, and based on findings from piloting the first complete set of lessons, we decided to include a second review lesson (Lesson 5).

Judgements of the consensus panel regarding the order in which the concepts should be taught were based on Wiggins’ and McTighe’s argument that a curriculum should be designed around the needs of learners trying to understand the big ideas and to perform the core tasks, rather than around the logic of the content. However, when designing and revising the lessons, we chose not to introduce the concepts in the order suggested by the consensus panel. One reason for this was that the logic underlying the organisation of the Key Concepts into three groups (claims, comparisons, and choices) corresponds directly to the needs of learners trying to understand the big ideas and to perform the core tasks (Table 4). Thus, it was both logical and consistent with learners’ needs to group the lessons into those three parts, as shown in Table 8. Another reason was that we felt it was important to include concepts from each of the three groups (claims, comparisons, and choices) in the lessons. Another explanation might be that the judgements of members of the consensus panel were not informed by experience teaching the concepts.

### What this study adds to what was previously known

When we prioritised concepts for the IHC primary school resources, the judgements were made by the research team. At that time, there were 32 Key Concepts in the framework.^15^ A network of teachers in Uganda assessed the relevance of the 32 concepts and determined that 24 of those concepts were relevant to primary school children in Uganda.^16^ However, all 24 concepts proved to be too much to learn in a school term. The early prototypes of the primary school resources that we created had too many concepts per lesson and took too long to teach in a normal school period (40 minutes). We also observed that the teachers needed time to repeat material from previous lessons. After each round of prototyping, we eliminated more Key Concepts from our list. We decided which ones to eliminate by considering the importance of the concepts and the difficulty that the children had learning them. The importance of the concepts was based on judgements made by members of the research team by:

- Each person individually identifying which of the 24 Key Concepts they considered most important
- Compilation and discussion of those judgements
- Voting on the concepts
- Reaching a consensus by informal discussion

The research team reached agreement that eight of the concepts were most important for our target population in Uganda. Three members of the research team also reviewed data from our piloting and user-testing and identified concepts that appeared to be too difficult to teach to 10 to 12-year-old children. Later, based on feedback from piloting the resources, we considered how the concepts were grouped in the lessons and the number of concepts being taught in each lesson. We ended up including 12 of the 24 concepts in the final version of the resources (Table 9) and reorganised them into three groups to simplify and clarify their purpose:

**Table 9.** Comparison of concepts in the IHC primary and secondary school resources

| Concept | Primary school resources | Secondary school resources |
| --- | --- | --- |
| Treatments can cause harms as well as benefits. | ✓ | ✓ |
| Large, dramatic effects are rare. |  | ✓ |
| Personal experiences or anecdotes alone are an unreliable basis for most claims. | ✓ | ✓ |
| Widely used treatments or those that have been used for decades are not necessarily beneficial or safe. | ✓ | ✓ |
| Treatments that are new or technologically impressive may not be better than available alternatives. | ✓ | ✓ |
| Opinions do not alone provide a reliable basis for deciding on the benefits and harms of treatments. | ✓ |  |
| Conflicting interests may result in misleading claims about the effects of treatments. | ✓ |  |
| Identifying effects of treatments depends on making comparisons. | ✓ | ✓ |
| Comparison groups should be as similar as possible. | ✓ | ✓ |
| If possible, people should not know which of the treatments being compared they are receiving | ✓ |  |
| Small studies may be misleading. | ✓ | ✓ |
| The results of single comparisons of treatments can be misleading | ✓ |  |
| Weigh the benefits and savings against the harms and costs of acting or not. | ✓ | ✓ |

- **Claims**: “questions you should ask when someone says something about a treatment”
- **Comparisons**: “questions that health researchers ask to find out more about the effects of treatments”
- **Choices**: “questions that you should ask when you are choosing whether to use a treatment”

The main reason for including fewer concepts in the secondary school resources was that 10 double (80-minute) periods were used for the primary school resources – twice as much time as for the secondary school resources. In addition, the primary school resources included printed materials for the students (including textbooks), whereas the secondary school resources are primarily digital resources for teachers, with optional handouts for students. Thus, they are more dependent on teachers being well-prepared to teach the lessons. The reason for using digital learning resources and limiting each lesson to a single 40-minute period was to increase the likelihood that use of the resources will be scaled up, if shown to be effective.

Other educational interventions to improve people’s ability to think critically about healthcare claims and decide what to do, and assessment tools, have included only a handful of the key concepts,^17^ and it is unclear how those concepts were prioritised.

### Strengths and limitations of this study

Strengths of this study include involvement of curriculum specialists, teachers, and researchers in the three countries for which the IHC Key Concepts were prioritised, use of explicit criteria, independent judgements by a panel of judges, and feedback from teachers and other stakeholders before finalising the priorities. Changes to the priorities based on feedback on prototypes and judgements made by the research team could be viewed as either a strength or a limitation. It is important to be pragmatic, and we view the changes made based on user-testing prototypes and pilot-testing all 10 lessons as essential. The nine concepts were not being taught in any of the three countries and teachers had no prior experience teaching the concepts.

An important limitation of this study is that the concepts were prioritised independently of the rest of the curriculum. Ideally, IHC Key Concepts could be prioritised together with other important concepts in the curriculum. Rwanda implemented a new competence-based curriculum in 2016. Uganda introduced its new competence-based curriculum for lower-secondary schools in 2020, and Kenya has plans to introduce a new competency-based curriculum by 2024.^18-20^ The new curricula in all three countries include critical thinking as a core competence and they include health topics. However, critical thinking about health is not explicitly included in any of the curricula, and both critical thinking and health are taught across subjects. This limited our ability to integrate the IHC Key Concepts into the curricula. In addition, teaching is exam-oriented in all three countries. Since IHC Key Concepts are currently not assessed in national examinations, this may further limit implementation of teaching the prioritised concepts.

An additional limitation is that we were not able to design resources to support a spiral curriculum, as originally intended. Ideally IHC Key Concepts would be introduced in primary school.^7^ What was learned in primary school could then be reinforced and built upon in secondary school over several school terms. We also did not prioritise competences or dispositions.^4^ However, these overlap substantially with the concepts, as reflected in the learning goals for the 10 lessons (Table 8).

### Implications

There is substantial overlap between the IHC Key Concepts that were prioritised for our primary school and secondary school resources. The included concepts appear to be a good starting point for teaching children and adolescents to think critically about health actions. There remains a need to develop a spiral curriculum and a series of resources that can be used in primary and secondary schools, and that can be easily adapted to different contexts.

## Conclusions

Both young people and adults are confronted with an excess of health information, including a large amount of misinformation. The ability to distinguish between reliable and unreliable information about the effects of health actions depends on understanding and applying concepts in the IHC Key Concept framework. Many people have not learned many of those concepts.^7, 21, 22^ It is not practical to teach or learn all the IHC Key Concepts in a single school term. Thus, is important to prioritise which concepts to teach. Using an iterative process with explicit criteria, we have prioritised nine concepts as a starting point for lower-secondary school students in Kenya, Rwanda, and Uganda.^3, 4^

## Data Availability

All relevant data are within the manuscript and its Supporting Information files.

## Acknowledgments

We are grateful to members of the teacher and student networks in Kenya, Rwanda, and Uganda who provided feedback on the results of the first consensus and feedback on prototypes of the lower-secondary-school resources, to members of the international and national advisory groups who provided feedback, and to other members of the research team for their input.

## Supporting information

S1 First consensus prioritisation of Key Concepts

S2 Second consensus prioritisation of Key Concepts

